# Deep learning based ischemic lesion markers on non-contrast head CT compared to CTP and DWI

**DOI:** 10.1101/2025.11.18.25340504

**Authors:** Henk van Voorst, Bin Jiang, Praneeta Konduri, Adrien ter Schiphorst, Aroosa Zamarud, Seena Dehkharghani, Lieselotte vandeWalle, Ewout Heylen, Yongkai Liu, Michael Mlynash, Soren Christensen, Nicole Yuen, Benjamin FJ Verhaaren, Abdelkader Mahammedi, Patrik Michel, Max Wintermark, Gregory W Albers, Greg Zaharchuk, Maarten G Lansberg, Jeremy J Heit

**Author notes:** **Disclosures** JJH is a consultant for Medtronic and Microvention. He is an advisory board member for MicroVention and iSchemaView. SD is a consultant and advisory board member for RAPID.AI and a consultant to Regeneron pharmaceuticals. He holds intellectual property and patents for electromagnetic instrumentation and algorithms unrelated to this work. BV is a member of a Data and Safety Monitoring Board for uniQure, outside the submitted work. AtS has nothing to disclose. PK is a cofounder and shareholder of inSteps BV and receives funds from 2025-26 NIH StrokeNet Training Program (U10NS086487).

## Abstract

**Background:** Quantification of ischemic brain tissue on non-contrast CT (NCCT) in acute ischemic stroke is challenging in the acute setting.

**Purpose:** To compare the spatial overlap and imaging marker agreement of acute ischemic regions of interest (ROIs) using deep-learning NCCT (DLNCCT) versus manual NCCT, CTP, and DWI-based ischemic segmentations.

**Methods:** We trained a deep learning model to segment ischemic ROIs using manual lesion annotations on admission NCCTs (DLNCCT). DLNCCT ischemic ROIs were compared with manual NCCT delineation, CTP (rCBF<30%/38%), and DWI within 5 hours after the NCCT or after recanalization in four external test sets. Spatial overlap was measured using the Dice Similarity Coefficient (DSC; mean±SD). For each ROI, we derived: average density (HU); modified net water uptake (mNWU in %); total volume (mL); and hypodense (<26HU) volume (mL), and assessed agreement via Bland–Altman (mean difference [95%CI]) and concordance correlation coefficient (CCC) analysis.

**Results:** 218 training (n=104/89/25 male/female/unknown, mean age 68±14 years) and 762 test cases (n=243/206/313 male/female/unknown, mean age 70±15 years) were used. Spatial overlap was 0.30±0.30 between DLNCCT and manual segmentation, 0.22±0.25 between DLNCCT and DWI, 0.10±0.19/0.14±0.21 between DLNCCT and CTP (rCBF<30%/<38%), and 0.15±0.22/0.21±0.24 between CTP (rCBF<30%/<38%) and DWI. DLNCCT vs. DWI mean differences of ischemic ROI derived imaging markers were -1HU (95%CI:-7;6) for average density (CCC:0.71), 4.9% (95%CI:-7.0;16.8) for mNWU (CCC:0.35), -16mL (95%CI:-108;76) for total volume (CCC:0.57), and -4mL (95%CI:-31;23) for hypodense lesion volume (CCC: 0.75).

**Conclusion:** Spatial overlap and agreement of imaging markers between DLNCCT and DWI ischemic ROIs were comparable to CTP and DWI.

**Summary Statement:** Ischemic injury on NCCT is identified and quantified by a deep-learning model with accuracy similar to CTP and DWI in stroke patients with a large vessel occlusion.

**Key results:**

- Deep-learning models can segment ischemic brain tissue on NCCT.
- Ischemic regions identified by our model demonstrate comparable overlap with ischemic core segmentation on CTP (Dice: 0.21±0.24) and DWI (Dice: 0.22±0.25).
- Deep learning NCCT showed high agreement with follow-up DWI in determining the hypodense (<26 HU) lesion volume (mean difference -4mL [95%CI:-31;23], CCC: 0.75).

## Introduction

Acute ischemic stroke (AIS) manifests primarily as regions of reduced attenuation on non-contrast CT (NCCT). Larger volumes and more severe hypodensity have been associated with malignant cerebral edema, hemorrhage, and worse outcomes (1–7). Volume and hypodensity imaging markers, such as net water uptake (NWU) (5,6), ischemic core volume (7), and severely hypodense (<26 HU) lesion volume (4), are associated with reduced benefits of intravenous thrombolysis (IVT), neuroprotective treatment, and endovascular treatment (EVT) (4–7). Therefore, volume and hypodensity quantification may facilitate targeted treatment decisions. Unfortunately, existing measures for hypodensity quantification in admission NCCT, such as the Alberta Stroke Program early CT score (ASPECTS), have poor inter-rater agreement and cannot accurately quantify volume or hypodensity (8).

Current methods to evaluate the volume and degree of hypodensity in ischemic brain tissue require segmentation on NCCT with input from advanced imaging techniques such as CT Perfusion (CTP) (9,10), post-treatment imaging (11), manual segmentation (4,6,12,13), or by comparison with a registered ASPECTS atlas (14). Each of these methods has shortcomings that prevent generalizable, easy, and accurate implementation in routine clinical practice. The development of deep learning methods to identify and quantify the volume and severity of hypodensity on baseline NCCT is of high interest, but prior attempts have suffered from insufficient performance and a lack of external validation (15–18).

We developed and externally validated a deep learning based ischemic region of interest (ROI) segmentation method using only baseline NCCT (DLNCCT). Our DLNCCT model measures volume and hypodensity severity in AIS patients with a large vessel occlusion. We compared the spatial overlap and agreement of imaging indices extracted from ischemic ROIs based on DLNCCT, manual delineations on NCCT, CTP ischemic core, and DWI lesion, hypothesizing that deep learning-based models would perform favorably in predicting the ischemic core in the acute setting.

## Methods

### Data

#### Training data

We trained an ischemic ROI segmentation model on 218 admission NCCTs from patients screened for the DEFUSE 3 trial, which evaluated endovascular treatment (EVT) for large-vessel occlusion (LVO) 6–16 hours after onset (19). To ensure a broad infarct size distribution, both enrolled patients and those excluded for large ischemic cores with admission CTP and NCCT were included. Three experts manually annotated ischemic lesions on NCCT without reference to other imaging except the infarcted hemisphere to reflect real-world uncertainty in lesion definition (15).

#### External test sets

Four datasets (AISD, ISLES24, Lausanne, CRISP2) were used for external validation of spatial and imaging-marker agreement (20–22) (Figure 1). Each included admission NCCT and manual or semi-automated lesion delineations on NCCT, CTP, or diffusion weighted imaging (DWI) after transfer or after treatment at follow-up (DWI). DWI follow-ups were included only for patients with excellent reperfusion (modified treatment in cerebral infarction score [mTICI]≥2C). Transfer DWIs were only included for patients with less than 5 hours between NCCT and DWI. More specific details for each dataset are described in Appendix A.

**Figure 1.**
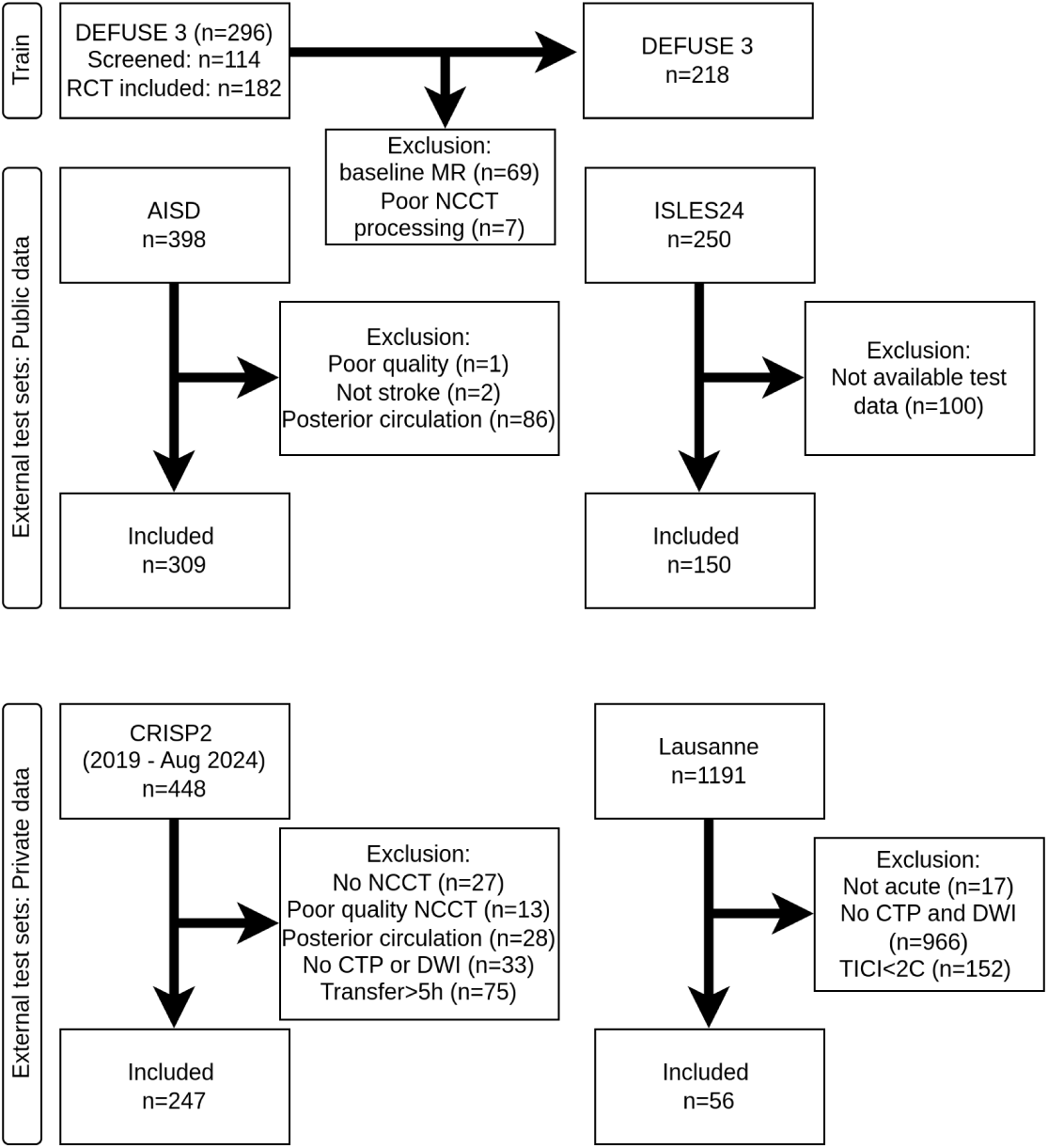
Train and external test datasets

### NCCT processing

We used the original admission NCCT (NCCT_original_), a mirrored NCCT (NCCT_mirrored_), and the ratio of the original and mirrored NCCT (Ratio = NCCT_original_ / NCCT_mirrored_) as a 3-channel input for training a 3D full-resolution no new UNet (nnUNet) (23). The NCCT_mirrored_ is obtained by flipping the NCCT_original_ in the lateral dimension and registering it back to the NCCT_original_ dimension using PyAnts’ translation, rigid, similarity, and double affine registration (TRSAA). The NCCT_mirrored_ and ratio image served to provide information on the density and relative density with respect to the contralateral non-ischemic hemisphere (Figure 2) (24). We trained for 1000 epochs, the best performing models were determined using the validation set. During training for each NCCT, one of the three raters’ lesion annotations was randomly selected as ground truth. We used a mean-ensemble of three models’ predictions, derived from subject-level 3-fold cross-validation during training, to generate the final predictions. We preferred a 3-fold over 5-fold ensemble as the results were comparable, but inference time was considerably shorter.

**Figure 2:**
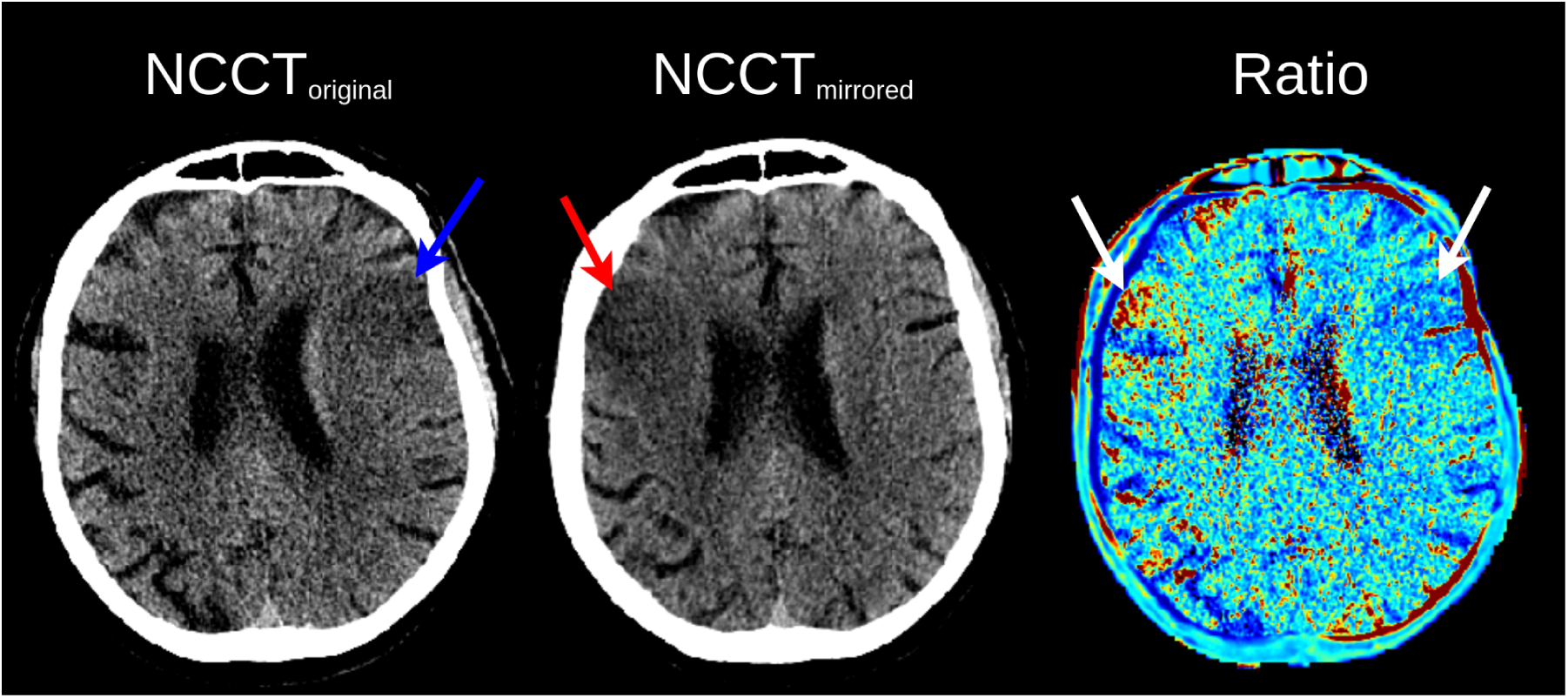
Input channels for the nnUNet. Left: Original admission NCCT (NCCT_original_) the blue arrow points to hypodensity. Middle: The mirrored NCCT (NCCT_mirrored_) can be obtained with a lateral flip and coregistration back to NCCT_original_; the red arrow points to the mirrored hypodensity. Right: Ratio image (Ratio) equals NCCT_original_ / NCCT_mirrored_ . Blue values in the ratio image (right white arrow) are below one and imply a percentage of HU depression compared to the contralateral non-affected hemisphere. Red values in the ratio image (left white arrow) imply a percentage increase in HU and are seen in the non-affected hemisphere.

### CTP and DWI processing

The first CT frame in the CTP sequence and the DWI were registered to the admission NCCT to enable ROI comparisons using simpleelastix 2.4.0 (CRISP2) and pyants 0.4.2 (AISD, Lausanne). All ISLES24 images were already aligned with the admission NCCT. Ischemic core masks based on a relative cerebral blood flow <30% or <38% (rCBF<30% or rCBF<38%) were extracted using RAPID 5.1.1 *(2020, iSchemaView)*. A previously developed and validated DWI lesion segmentation algorithm was used to segment the ischemic ROI in DWI (25).

### Evaluation measures and statistical analyses

Volume and density measurements on admission NCCT were computed for each of the ischemic ROIs as derived from i. DLNCCT; ii. manual annotations of NCCT (NCCT-manual; only available for AISD), iii. CTP-based ischemic core at prescribed perfusion thresholds CTP(rCBF<30%) and CTP(rCBF<38%); and iv. semi-automatically segmented DWI lesions where available. All ischemic ROIs were constrained to areas within a brainmask extracted using totalsegmentator 2.4.0, in the infarcted hemisphere, excluding regions not covered by the CTP scan (for comparisons to CTP only) or in the cerebellum or pons. Imaging markers extracted from each ROI included: average density (HU), net water uptake (NWU = 100*(1-average HU lesion / average HU lesion location NCCT_mirrored_), total lesion volume, and severely hypodense(<26 HU) volume. Per Minnerup et al., NWU calculations excluded voxels <20 or >80 HU to avoid bone and cerebospinal fluid artifacts (9). We refer to NWU in this manuscript as modified NWU (mNWU), as we did not use a low cerebral blood flow mask from perfusion CT for voxel selection as originally described (9).

To evaluate spatial overlap between predicted and ground truth reference segmentations, we report the dice similarity coefficient (DSC), true positive rate (TPR), positive predictive value (PPV), absolute volume difference (AVD), and average Hausdorff distance (AHD). We separately report spatial overlap measures for a subgroup of patients with a DWI or manually segmented NCCT lesion exceeding 50 mL, to emphasize performance in a subpopulation with large lesion volumes for which poor prognostication using imaging markers may affect treatment decisions. Dataset-specific results appear in Appendix B.

Agreement between ischemic ROI methods for imaging marker extraction was assessed with Bland–Altman analyses (mean difference, 95%CI) and the concordance correlation coefficient (CCC). For non-normal distributions and heteroskedasticity, we scaled the variables using log_10_ transform for visualization and used robust standard errors for regression analyses. Since additional clinical variables may inform variations in estimation errors of imaging markers between ischemic ROI methods, we used conditional Bland-Altman analyses and report the mean explained difference with 95% CI using a regression model (26), to evaluate whether the difference between DLNCCT and DWI ischemic ROI methods for imaging marker measurements can be explained by the admission National Institutes of Health Stroke Scale (NIHSS), onset-to-NCCT time, ASPECTS, and Tan collateral score.

## Results

Table 1 shows baseline characteristics across datasets. Although patients were only included if admission NCCT was available (AISD: 309, ISLES: 150, Lausanne: 56, CRISP2: 247), the availability of concomitant CTP (AISD: 0, ISLES24: 150, Lausanne: 38, CRISP2: 200) and DWI (AISD: 309, ISLES24: 150, Lausanne: 56, CRISP2: 247) varied across datasets.

**Table 1:** Baseline Characteristics of all datasets.

|  | **DEFUSE3**<br>(n=218) | **AISD**<br>(n=309) | **ISLES24**<br>(n=150) | **Lausanne**<br>(n=56) | **CRISP2**<br>(n=247) |
| --- | --- | --- | --- | --- | --- |
| **Age in years**<br>median(IQR) | 69 (58;79) | Not<br>available* | 75 (64;82) | 65 (54;76) | 73 (61;82) |
| **Sex**<br>Male/Female | 104/89<br>(54%/46%) | Not<br>available* | 69/78<br>(47%/53%) | 32/24<br>(57%/43%) | 142/104<br>(58%/42%) |
| **Onset or last**<br>**known well to**<br>**NCCT time in**<br>**hours**<br>median(IQR) | 10 (8;12) | Not available<br>(<24 hours)* | 2 (1;3) | 3 (2;10) | 3 (1;9)* |
| **Admission**<br>**NIHSS**<br>median(IQR) | 16 (12;21) | Not<br>available* | 10 (6;17) | 8 (4;17) | 14 (9;20) |

|  | **DEFUSE3**<br>**(n=218)** | **AISD**<br>**(n=309)** | **ISLES24**<br>**(n=150)** | **Lausanne**<br>**(n=56)** | **CRISP2**<br>**(n=247)** |
| --- | --- | --- | --- | --- | --- |
| **ASPECTS**<br>median(IQR) | 8 (7;9) | Not<br>available* | 8 (6;10) | 9 (7;10) | 9 (8;10) |
| **CTA Tan**<br>**collateral score**<br>Count per<br>subscore |  | Not<br>available* |  |  |  |
| **0** | 35 (32.1%)** |  | 4 (2.7%) | 5 (9.1%) | 7 (2.9%) |
| **1** |  |  | 42 (28.2%) | 16 (29.1%) | 62 (25.6%) |
| **2** | 74 (67.9%)** |  | 70 (47.0%) | 18 (32.7%) | 106 (43.8%) |
| **3** |  |  | 33 (22.1%) | 16 (29.1%) | 67 (27.7%) |
| **DLNCCT**<br>**volume in mL**<br>median(IQR) | 14 (5;35) | 10 (2;41) | 9 (1;27) | 4 (1;17) | 5 (1;12) |
| **CTP ischemic**<br>**core volume in**<br>**mL (rCBF<30%)**<br>median(IQR) | 11 (2;38) | Not<br>available* | 4 (0;20) | 0 (0;8) | 0 (0;14) |
| **DWI lesion**<br>**volume in mL**<br>median(IQR) | Not<br>available** | 18 (7;58) | 9 (2;31) | 14 (2;44) | 15 (4;35) |

\*: For AISD, no clinical variables and CTP were available. \*\*: In DEFUSE 3, collaterals were scored as below or above 50% (combining Tan scores 0-1 and 2-3), and DWI was not available.

### Spatial overlap between ischemic ROIs

Spatial overlap measures were compared for all ischemic ROI methods across all subjects and for a subgroup with large infarcts (>50mL) (Table 2). DLNCCT achieved the highest spatial overlap with manual NCCT segmentations (DSC all subjects: 0.30 ±0.30, >50mL: 0.50 ±0.29), which was comparable to cross-validation performance (DSC all subjects: 0.38 ±0.26, >50mL: 0.67 ±0.14) and interrater spatial overlap (DSC all subjects: 0.32 ±0.23, >50mL: 0.58 ±0.19).

**Table 2:** Spatial overlap between ischemic region of interest (ROI) segmentation methods.

| Ischemic ROI comparison | Number of subjects in analysis | | Dice Similarity Coefficient (mean±SD) | | True positive rate (sensitivity) (mean±SD) | | Positive predictive value (precision) (mean±SD) | | Absolute volume difference (AVD) (mean±SD) | | Average Hausdorff Distance (mm) (mean±SD) | |
| --- | --- | --- | --- | --- | --- | --- | --- | --- | --- | --- | --- | --- |
|  | all | >50mL | all | >50mL | all | >50mL | all | >50mL | all | >50mL | all | >50mL |
| Interrater Manual-manual (training data)* | 218 | 32 | 0.32 ±0.23 | 0.58 ±0.19 | 0.28 ±0.22 | 0.54 ±0.21 | 0.50 ±0.31 | 0.76 ±0.15 | 17 ±21 | 49 ±31 | 7 ±9 | 3 ±3 |
| DLNCCT-Manual (cross-validation)** | 218 | 32 | 0.38 ±0.26 | 0.67 ±0.14 | 0.48 ±0.32 | 0.73 ±0.16 | 0.39 ±0.26 | 0.69 ±0.12 | 13 ±14 | 34 ±17 | 8 ±13 | 2 ±2 |
| DLNCCT-Manual*** | 309 | 105 | 0.30 ±0.30 | 0.50 ±0.29 | 0.30 ±0.30 | 0.47 ±0.29 | 0.43 ±0.38 | 0.66 ±0.32 | 25 ±38 | 46 ±51 | 15 ±16 | 7 ±11 |
| DLNCCT-DWI | 762 | 198 | 0.22 ±0.25 | 0.37 ±0.27 | 0.21 ±0.25 | 0.32 ±0.26 | 0.34 ±0.36 | 0.64 ±0.33 | 27 ±41 | 71 ±57 | 16 ±18 | 8 ±9 |
| DLNCCT-CTP (rCBF<38%) | 388 | 78 | 0.14 ±0.21 | 0.30 ±0.27 | 0.16 ±0.26 | 0.34 ±0.33 | 0.22 ±0.29 | 0.45 ±0.32 | 19 ±25 | 39 ±34 | 15 ±16 | 9 ±11 |
| DLNCCT-CTP (rCBF<30%) | 388 | 78 | 0.10 ±0.19 | 0.24 ±0.26 | 0.13 ±0.26 | 0.32 ±0.35 | 0.14 ±0.24 | 0.32 ±0.29 | 15 ±22 | 32 ±30 | 18 ±22 | 9 ±9 |
| CTP(rCBF<38%) - DWI | 388 | 78 | 0.21 ±0.24 | 0.49 ±0.21 | 0.25 ±0.26 | 0.43 ±0.23 | 0.28 ±0.33 | 0.72 ±0.24 | 20 ±24 | 47 ±33 | 10 ±15 | 4 ±9 |
| CTP(rCBF<30%) - DWI | 388 | 78 | 0.15 ±0.22 | 0.38 ±0.24 | 0.14 ±0.21 | 0.31 ±0.23 | 0.24 ±0.34 | 0.67 ±0.34 | 20 ±27 | 59 ±37 | 14 ±23 | 4 ±5 |

\*: Mean and SD represent the mean of raters (mean of raters 1 vs 2, 1 vs 3, and 2 vs 3) distributed across DEFUSE 3 subjects. \*\*: cross-validation results represent mean and SD by pooling all validation sets and taking the mean performance metric across three raters (mean of DLNCCT vs rater 1, DLNCCT vs rater 2, DLNCCT vs rater 3) across subjects. \*\*\*: AISD was the only dataset with a manual NCCT ground-truth segmentation per subject.

DLNCCT overlap was higher when compared to DWI (DSC all subjects: 0.22 ±0.25, >50mL: 0.37 ±0.27) than when DLNCCT was compared to the ischemic core on CTP using rCBF<30% (DSC all subjects: 0.10 ±0.19, >50mL: 0.24 ±0.26) and rCBF<38% (all subjects: 0.14 ±0.21, >50mL: 0.30 ±0.27). The overlap between the CTP ischemic core and DWI lesion showed worse performance for rCBF<30% (all subjects: 0.15 ±0.22, >50mL: 0.38 ±0.24), and similar performance for rCBF<38% (all subjects: 0.21 ±0.24, >50mL: 0.49 ±0.21) to spatial correspondences between DLNCCT and DWI. Low spatial agreement of the DLNCCT method was predominantly due to underestimation (lower TPR) compared to oversegmentation (moderate PPV). The average Hausdorff Distance was comparable across ROI methods. Spatial overlap between DLNCCT and DWI ROIs for each dataset separately is presented in Appendix Table 1. DSC was comparable between AISD (DSC all subjects: 0.24±0.26; >50 mL: 0.40±0.26), ISLES24 (DSC all subjects: 0.19±0.23; >50 mL: 0.44±0.25), Lausanne (DSC all subjects: 0.22±0.24; >50 mL: 0.36±0.29), but not for large infarcts in CRISP2 (DSC all subjects: 0.21±0.24; >50 mL: 0.28±0.28). Figure 3 shows example NCCT and DWI pairs from each dataset. Despite considerable variability in image quality between datasets, spatial and imaging marker agreement were comparable (Appendix Table 1). Differences between NCCT, CTP, and DWI ROIs may be due to infarct expansion (Figure 3 row 1 and row 2) or subtle misalignment (Figure 3 row 4), which can be observed by comparing the ventricles.

**Figure 3.**
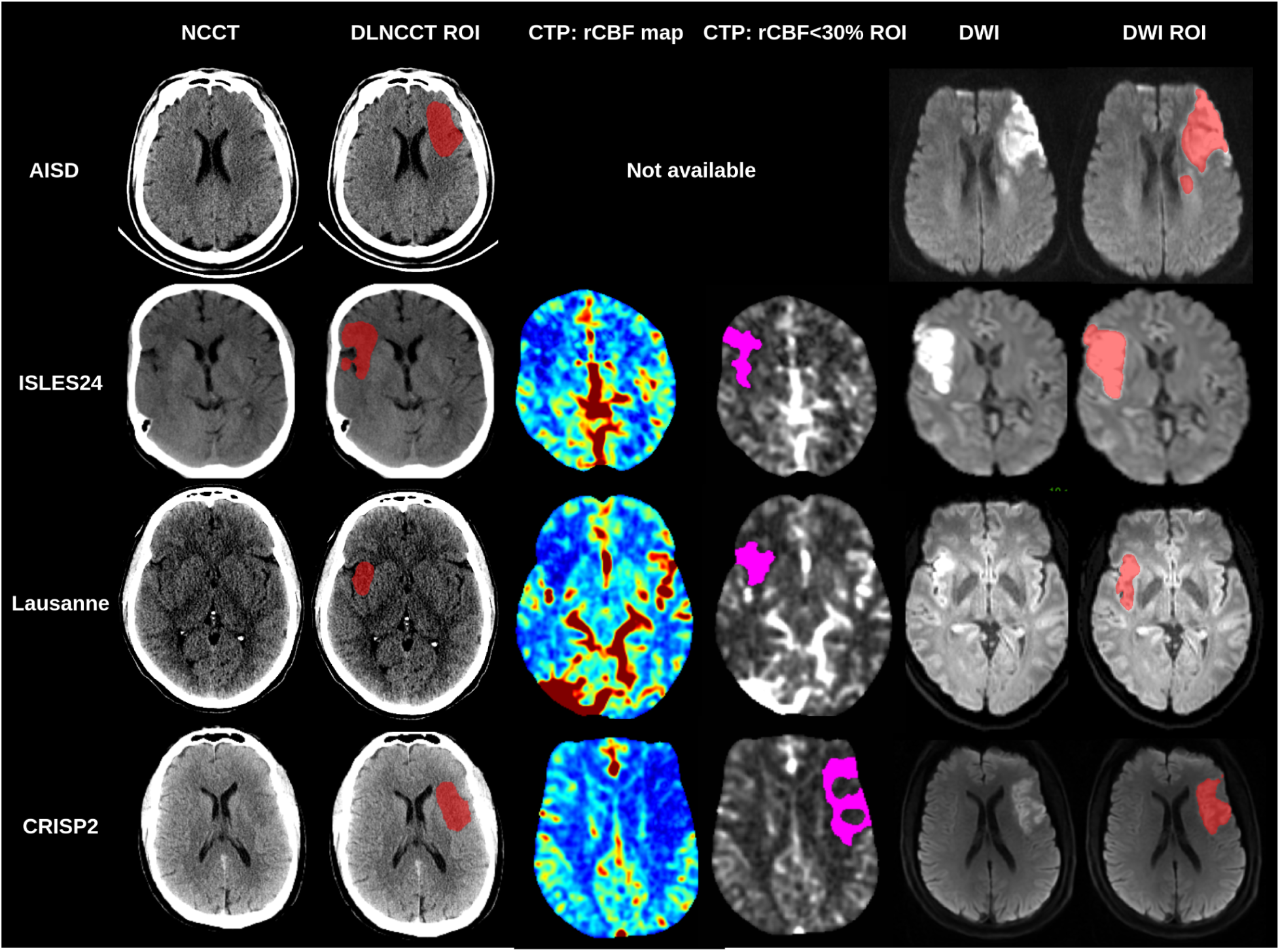
Example NCCT and DWI pairs with ischemic region of interest (ROI) segmentation from each dataset included. For AISD, no CTP imaging was available.

### Imaging marker agreement between ischemic ROIs

We performed Blant-Altman analyses and CCC to compare ischemic ROIs across imaging techniques (Table 3 and Figure 4). Similar to metrics describing spatial overlap, imaging markers extracted with DLNCCT were most comparable to those extracted from manual NCCT ROIs, followed by those from manual DWI and CTP ischemic ROIs. Mean differences for average lesion density, mNWU, and hypodense lesion volume were close to zero, but the 95%CI showed considerable differences in imaging marker values between ischemic ROI methods when considering the variability of these imaging markers (Table 1). DLNCCT slightly underestimated total and hypodense lesion volume compared to manual NCCT, DWI, and CTP (rCBF<38%) ROIs, and overestimated compared to CTP(rCBF<30%). CCC was higher for hypodense lesion volume than for total lesion volume and was lower for average density and mNWU. In Appendix Table 2, we describe the imaging markers agreement between DLNCCT and DWI ischemic ROIs across datasets.

**Figure 4:**
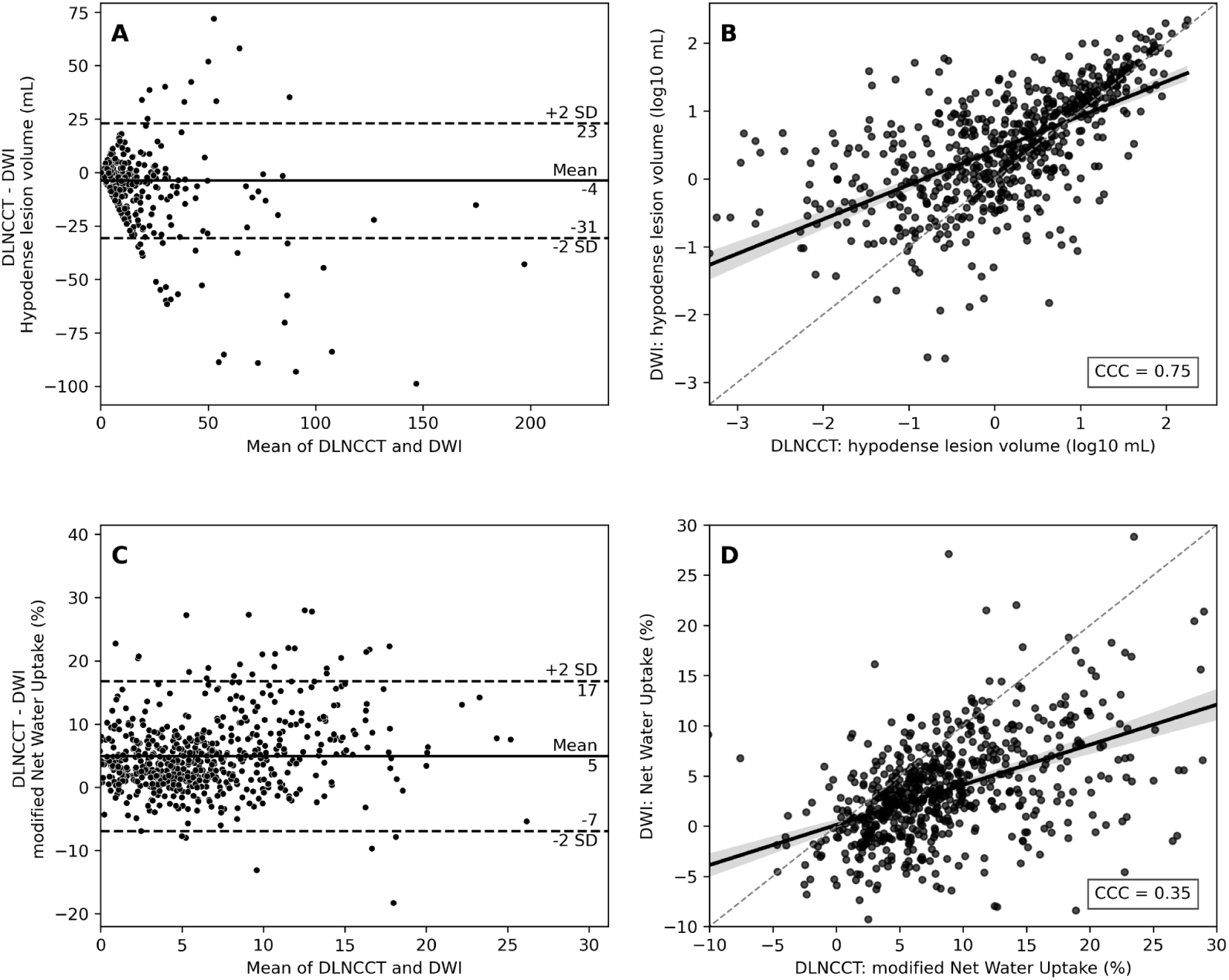
Agreement of imaging marker for DLNCCT vs. DWI-based ischemic ROIs. A/C: Bland-Altman analyses of hypodense (<26HU) lesion volume (A) and modified Net Water Uptake (C), positive values indicate DLNCCT overestimates compared to DWI. B/D: Concordance Correlation Coefficient (CCC) plot of logarithmic scaled hypodense (<26HU) lesion volume (B) and modified Net Water Uptake (D) between DLNCCT (x-axis) and DWI (y-axis) ischemic ROIs.

**Table 3.** Agreement of imaging markers between varying ischemic region of interest segmentation methods.

| Ischemic ROI comparison | Average lesion density (HU) |  | modified Net Water Uptake (%) |  | Total lesion volume (mL) |  | Hypodense (<26 HU) lesion volume (mL) |  |
| --- | --- | --- | --- | --- | --- | --- | --- | --- |
|  | Mean diff (95%CI) | CCC | Mean diff (95%CI) | CCC | Mean diff (95%CI) | CCC | Mean diff (95%CI) | CCC |
| DLNCCT-Manual (cross-validation)* | 0 (-4;4) | 0.86 | 0.3 (-8.1;8.7) | 0.62 | 2 (-25;30) | 0.92 | 1 (-7;8) | 0.95 |
| DLNCCT-Manual** | 1 (-5;7) | 0.59 | 0.2 (-11.1;11.4) | 0.62 | -11 (-97;75) | 0.72 | -6 (-35;23) | 0.83 |
| DLNCCT-DWI | -1 (-7;6) | 0.71 | 4.9 (-7.0;16.8) | 0.35 | -16 (-108;76) | 0.57 | -4 (-31;23) | 0.75 |
| DLNCCT-CTP (rCBF<38%) | 0 (-7;7) | 0.75 | 3.4 (-7.6;14.4) | 0.35 | -9 (-68;51) | 0.44 | -2 (-25;21) | 0.59 |
| DLNCCT-CTP (rCBF<30%) | 0 (-9;10) | 0.62 | 3.4 (-12.0;18.9) | 0.25 | 2 (-49;53) | 0.46 | 1 (-20;22) | 0.58 |
| CTP(rCBF<38%) - DWI | -1 (-8;6) | 0.75 | 0.7 (-10.7;12.0) | 0.46 | -4 (-64;55) | 0.65 | -1 (-27;25) | 0.64 |
| CTP(rCBF<30%) - DWI | -1 (-11;9) | 0.66 | 0.4 (-14.3;15.1) | 0.37 | -15 (-73;44) | 0.55 | -4 (-29;20) | 0.59 |

Mean difference (ROI1-ROI2) and 95%CI of Bland-Altman analysis. CCC: Spearman Concordance correlation coefficient. \*: Cross-validation results from the training dataset (DEFUSE 3), per subject, the mean performance metric compared of three raters was used. \*\*: AISD was the only dataset with a manual NCCT ground-truth segmentation per subject.

### Explained variation between DLNCCT and DWI-based imaging markers

To better understand how clinical variables influence and may guide DLNCCT ischemia assessment compared to DWI and to identify subgroups with greater agreement, we performed conditional Bland-Altman analyses (Table 4). More severe stroke symptoms, as determined with the admission NIHSS, were significantly associated with underestimation of DLNCCT total lesion volume (-0.38mL [95%CI:-0.71;-0.04], p=0.03) and hypodense lesion volume (-0.13mL [95%CI:-0.25;0.0], p=0.04). Baseline ASPECTS was associated with DLNCCT overestimation of average lesion density (0.15HU [95%CI:0.03;0.28], p=0.02) and NWU (0.37% [95%CI:0.12;0.63], p=0.005), underestimation of total lesion volume (-5.38 mL [95%CI:-7.96;-2.79], p<0.001), and hypodense lesion volume (-1.15mL [95%CI:-2.04;-0.27], p=0.01) (Figure 5A-B). Moreover, a per point increase of the CTA Tan collateral score was observed to overstimate DLNCCT total lesion volume (4.89mL [95%CI:1.21;8.56], p=0.009) and hypodense lesion volume (1.67mL [95%CI:0.54;2.8], p=0.004). All other comparisons were not statistically significant (Figure 5C-D).

**Figure 5.**
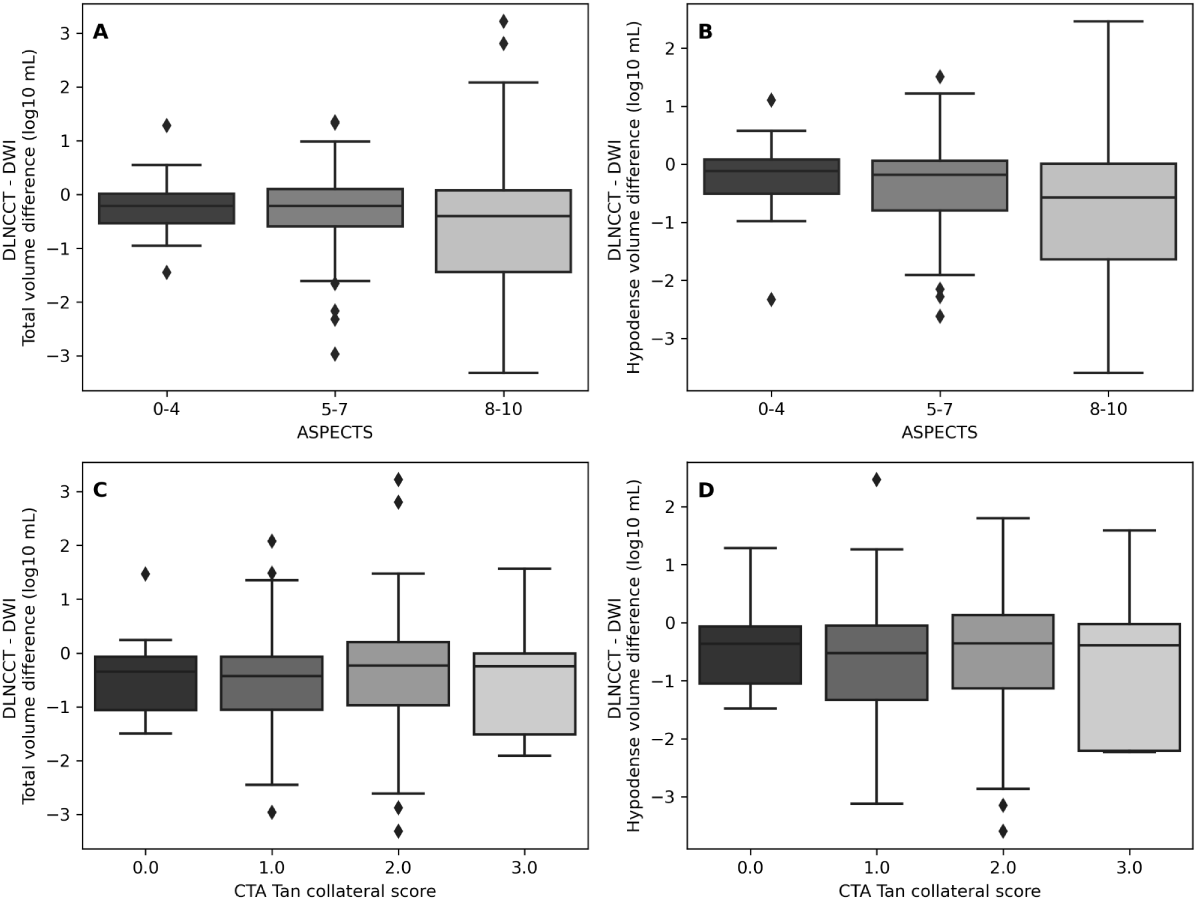
Conditional Bland-Altman analyses. A/B: DLNCCT - DWI vs ASPECTS. C/D: DLNCCT - DWI vs CTA Tan collateral score. A/C: Total lesion volume difference. B/D: Hypodense (<26HU) lesion volume difference. Log10 transformation was applied to volume measures before subtraction (log10(DLNCCT volume) - log10(DWI volume)).

**Table 4.** Conditional Bland-Altman Analyses for DLNCCT vs. DWI-based ischemic ROIs. Positive values indicate more than proportionate overestimation of DLNCCT compared to DWI per unit increase of the baseline variable. *: p<0.05, **:p<0.01, ***:p<0.001.

| Baseline variable | Average lesion density (HU)<br>Mean difference (95%CI) | Net Water Uptake (%)<br>Mean difference (95%CI) | Total lesion volume (mL)<br>Mean difference (95%CI) | Hypodense (<26 HU) lesion volume (mL)<br>Mean difference (95%CI) |
| --- | --- | --- | --- | --- |
| Admission NIHSS | -0.01 (-0.05;0.04) | 0.01 (-0.07;0.10) | -0.38 (-0.71;-0.04)* | -0.13 (-0.25;0.0)* |
| Onset or last known well to NCCT time (hours) | -0.03 (-0.07;0.02) | -0.02 (-0.10;0.06) | 0.37 (-0.05;0.79) | 0.04 (-0.13;0.21) |
| Baseline ASPECTS | 0.15 (0.03;0.28)* | 0.37 (0.12;0.63)* | -5.38 (-7.96;-2.79)*** | -1.15 (-2.04;-0.27)* |
| CTA Tan Collateral score | 0.19 (-0.16;0.54) | 0.33 (-0.29;0.95) | 4.89 (1.21;8.56)** | 1.67 (0.54;2.8)** |

## Discussion

In this study, we developed and validated a deep learning model to segment and quantify hypodense ischemic brain tissue on NCCT in acute ischemic stroke patients with large vessel occlusions. Despite suboptimal spatial overlap for smaller lesions, imaging marker extraction using our DLNCCT method was comparable to manually annotated, CTP, and DWI-based ischemic ROIs.

Automated segmentation of ischemic brain tissue on NCCT is highly desirable, given the large use of NCCT in stroke patient evaluation. However, accurate identification and quantification of the ischemic core on NCCT is notoriously challenging, subject to high inter-rater disagreement, and affected by the time since stroke onset (8,15). These inherent limitations in NCCT evaluation have led to the common use of advanced imaging techniques, such as CTP, which more accurately measure ischemic brain tissue (10). Due to the limited availability of CTP, a deep learning model that can identify ischemic brain tissue on NCCT alone, with results comparable to CTP or DWI, is highly desirable.

Several studies have explored deep-learning-based ischemia segmentation on presentation NCCT (15–18,27). Gauriau et al. performed the largest study to date, training on admission NCCT with ≤3-hour DWI as the reference standard and demonstrating strong internal performance for lesion detection and volume estimation (18). Ischemia segmentation using automated or deep learning methods is quite variable in what is measured and in performance (15–18,27). Most prior studies have focused on lesion volume measurement, lacked multimodal segmentation comparisons, and omitted external validation despite the availability of public datasets (15–18,27). Liang et al. and our study present the only two datasets with manual admission NCCT annotations as ground truth for training segmentation models (20). Deep learning methods trained with registered CTP ischemic core (17,27) or DWI diffusion restriction (18) as ground truth may yield different ischemic ROIs on NCCT. Moreover, NCCT hypodensity does not always align with the ischemic core and represents a smaller fraction of the DWI lesion when considerable time has passed (28), introducing variation in the segmentation, imaging marker estimates, and outcome associations. Variations in modalities for obtaining training data ground truths may also complicate comparisons of methods, as in-domain comparisons are likely to yield higher performance metrics. The use of multimodal segmentation comparison and the overall favorable agreement of the DLNCCT method with established measures of the ischemic core in our study are therefore notable.

In our study, automated DLNCCT segmentation and manual NCCT delineation had imperfect spatial agreement, but density and hypodense volume imaging markers estimates were comparable with underestimated volumes and slightly overestimated density measures. CTP and DLNCCT-based ROIs showed low spatial overlap with each other (Table 2: row DLNCCT-CTP) but exhibited comparable differences relative to DWI ROIs (Table 2: row DLNCCT-DWI vs. CTP-DWI), which may indicate accurate DLNCCT estimations. Our DLNCCT method shows poorer spatial overlap and correspondence between imaging markers for smaller lesions. Although the DSC is inherently worse for small lesions, another possible explanation for this finding is that patients with smaller ischemic cores may be more likely to have been imaged in earlier time windows, when ischemic injury is less conspicuous on NCCT. In support of these hypotheses, we observed improved agreement for hypodense volume segmentation in patients with lower ASPECTS and poor collateral scores. Although this limited overlap for small lesions may partially limit model performance, prognostication and treatment effect estimation using imaging markers in patients with large hypodense lesions may be more relevant for medical decision-making. Therefore, our model performs favorably when compared to prior studies that have addressed this challenging imaging problem (15–18,20).

The validity and reproducibility of hypodense lesion volume segmentation using DLNCCT has considerable implications for clinical practice, and DLNCCT may be a useful tool for patient prognostication and treatment decisions. Yogendrakumar et al. established a hypodense lesion volume larger than 26 mL, determined from manual ischemic ROI segmentations on admission NCCT, as the first imaging marker associated with an absence of endovascular treatment benefit in a post-hoc analysis of the SELECT 2 trial (4). Findings from our study indicate that a combined volume-density estimate, such as volume with ≤26HU, yielded more consistent values across ischemic ROI methods than total volume or NWU. This may partially explain the stronger association of hypodense lesion volume with treatment effect than NWU (4,6). Differences in ischemic ROI definitions likely account for discrepancies in mNWU. The original NWU approach used a low–CBV region within the CTP core (9), whereas DLNCCT defines a broader ischemic ROI. Such global ROIs may include fewer hypodense voxels, leading to inaccurate mNWU estimates. Thus, mNWU may not be optimal for the DLNCCT method.

Our study has several limitations. In the absence of an absolute ground truth, we cross-compared ischemic ROI methods. Hence, conclusions can only be drawn for each method with respect to another. Ideally, ROI comparisons would be based on CTP and DWI imaging acquired with minimal delay from the admission NCCT as the ischemic lesion grows. In the acute ischemic stroke setting, this may interfere with treatment and is often considered impractical. Due to inherent imaging delays in our analyses, spatial overlap and imaging markers may also vary across different acquisition times. Delays between admission NCCT and DWI, have likely resulted in a larger underestimation of volume and overestimation of density measures for our DLNCCT method, as DWI ROIs are larger and include non-ischemic voxels in NCCT. Future studies should compare imaging markers computed using different methods to prognosticate outcomes and estimate the heterogeneity of treatment effect. For treatments with lower benefits, such as thrombolysis and neuroprotective treatments in patient populations in extended time windows or with larger lesions, using admission NCCT imaging markers may be more accurate and valuable for treatment decisions. Since we trained and evaluated the deep learning method only for acute ischemic stroke cases, further research is needed to determine if the proposed method can also be used to diagnose or rule out ischemic stroke.

## Conclusion

Deep learning-based segmentation of an ischemic ROI on admission NCCT and CTP ischemic core segmentations have comparable spatial agreement with DWI ischemic lesion segmentation. DLNCCT shows relatively small mean differences in volume and hypodensity measurements compared with CTP and DWI ischemic ROIs, yet substantial variability persists despite this overall agreement.

## Data Availability

All data produced in the present study are available upon reasonable request to the authors

## Abbreviations

AHD: Average Hausdorff Distance
ASPECTS: Alberta Stroke Program early CT score
CCC: Concordance correlation coefficient
CTP: CT perfusion
DLNCCT: Deep learning based NCCT ischemic ROI segmentation
DSC: Dice Similarity Coefficient
DWI: Diffusion Weighted Imaging
EVT: Endovascular treatment
LVO: Large vessel occlusion
mTICI: modified treatment in cerebral infarction score
NCCT: noncontrast CT
mNWU: modified Net Water Uptake
PPV: Positive predictive value (equivalent to precision)
rCBF: Relative Cerebral Blood Flow
ROI: region of interest
SD: Standard deviation
TPR: True positive rate (equivalent to sensitivity)

## Appendix

### Appendix A: Datasets

More specific details for each data set include:

- AISD: A public dataset of anterior-circulation strokes with baseline NCCT and DWI within 24 hours. Posterior strokes were excluded. Recanalization rates and clinical data were not available. This was the only dataset with manual ischemic ROI segmentation on admission NCCT.
- ISLES24: The Ischemic Stroke Lesion Segmentation Challenge 2024 (ISLES24) is a public dataset for a challenge that was aimed at predicting follow-up DWI lesion segmentations using admission NCCT and CTP for patients with an LVO. Follow-up DWI was acquired 2-9 days after baseline imaging for ISLES24; all patients in this dataset achieved mTICI≥2C after EVT. Clinical baseline and outcome variables are publicly available.
- Lausanne Registry: Patients from the Swiss national registry for monitoring endovascular treatment efficacy and safety were included if baseline NCCT and CTP, and follow-up DWI were available after achieving mTICI≥2C post-EVT.
- CRISP2: The prospective CTP to predict Response to Recanalization in Ischemic Stroke Project (CRISP2) is an ongoing registry that includes patients with LVOs who are transferred from a primary to a secondary stroke center for EVT. Primary stroke center admission imaging included baseline NCCT and CTP, and DWI after transfer to a comprehensive stroke center. We used the data partition that includes patients up to August 2024. Patients were only included if the time between baseline imaging and DWI in the comprehensive stroke center was less than 5 hours.

### Appendix B supplemental results

**Appendix Table 1:** Spatial overlap between Deep Learning based noncontrast CT (DLNCCT) and Diffusion Weighted Imaging (DWI) based ischemic regions of interest across all used datasets.

| Datasets comparing DLNCCT-DWI ischemic ROIs | Number of subjects in analysis |  | Dice Similarity Coefficient (mean±SD) |  | True positive rate (sensitivity) (mean±SD) |  | Positive predictive value (precision) (mean±SD) |  | Absolute volume difference (AVD) (mean±SD) |  | Average Hausdorff Distance (mm) (mean±SD) |  |
| --- | --- | --- | --- | --- | --- | --- | --- | --- | --- | --- | --- | --- |
|  | all | >50mL | all | >50mL | all | >50mL | all | >50mL | all | >50mL | all | >50mL |
| AISD | 309 | 105 | 0.24 ±0.26 | 0.40 ±0.26 | 0.22 ±0.24 | 0.35 ±0.25 | 0.37 ±0.37 | 0.62 ±0.33 | 34 ±50 | 73 ±65 | 16 ±17 | 8 ±11 |
| ISLES24 | 150 | 24 | 0.19 ±0.23 | 0.44 ±0.25 | 0.22 ±0.28 | 0.40 ±0.28 | 0.27 ±0.32 | 0.66 ±0.26 | 21 ±29 | 61 ±39 | 19 ±22 | 5 ±6 |
| Lausanne | 56 | 15 | 0.22 ±0.24 | 0.36 ±0.29 | 0.23 ±0.28 | 0.33 ±0.3 | 0.35 ±0.35 | 0.62 ±0.35 | 22 ±32 | 60 ±42 | 15 ±19 | 6 ±6 |
| CRISP2 | 247 | 54 | 0.21 ±0.24 | 0.28 ±0.28 | 0.19 ±0.23 | 0.22 ±0.23 | 0.35 ±0.36 | 0.67 ±0.34 | 23 ±37 | 75 ±51 | 13 ±16 | 9 ±8 |

**Appendix Table 2:**
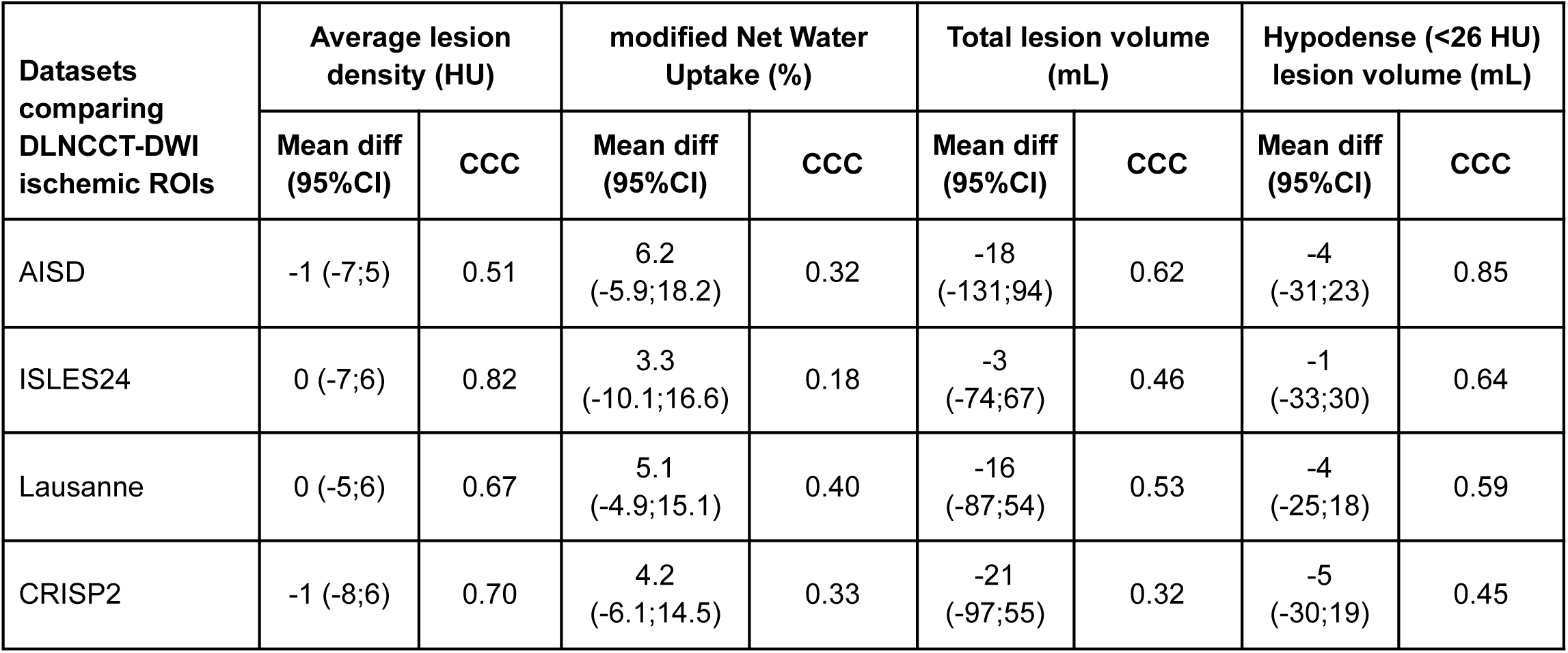
Imaging marker agreement between DLNCCT and DWI ischemic region of interest (ROI) segmentations. Mean difference (DLNCCT-DWI) and 95%CI of Blant-Altman analysis. Spearman Concordance correlation coefficient (CCC).

| Datasets comparing DLNCCT-DWI ischemic ROIs | Average lesion density (HU) |  | modified Net Water Uptake (%) |  | Total lesion volume (mL) |  | Hypodense (<26 HU) lesion volume (mL) |  |
| --- | --- | --- | --- | --- | --- | --- | --- | --- |
|  | Mean diff (95%CI) | CCC | Mean diff (95%CI) | CCC | Mean diff (95%CI) | CCC | Mean diff (95%CI) | CCC |
| AISD | -1 (-7;5) | 0.51 | 6.2 (-5.9;18.2) | 0.32 | -18 (-131;94) | 0.62 | -4 (-31;23) | 0.85 |
| ISLES24 | 0 (-7;6) | 0.82 | 3.3 (-10.1;16.6) | 0.18 | -3 (-74;67) | 0.46 | -1 (-33;30) | 0.64 |
| Lausanne | 0 (-5;6) | 0.67 | 5.1 (-4.9;15.1) | 0.40 | -16 (-87;54) | 0.53 | -4 (-25;18) | 0.59 |
| CRISP2 | -1 (-8;6) | 0.70 | 4.2 (-6.1;14.5) | 0.33 | -21 (-97;55) | 0.32 | -5 (-30;19) | 0.45 |

